# Predictors of Clinical Outcomes among People with HIV and Tuberculosis Symptoms after Rapid Treatment Initiation in Haiti

**DOI:** 10.1101/2024.06.19.24309189

**Authors:** Aaron Richterman, Nancy Dorvil, Vanessa Rivera, Heejung Bang, Patrice Severe, Kerylyne Lavoile, Samuel Pierre, Alexandra Apollon, Emelyne Dumond, Guyrlaine Pierre Louis Forestal, Vanessa Rouzier, Patrice Joseph, Pierre-Yves Cremieux, Jean W Pape, Serena P Koenig

**Affiliations:** University of Pennsylvania Perelman School of Medicine, Philadelphia, Pennsylvania, United States of America; Haitian Group for the Study of Kaposi’s Sarcoma and Opportunistic Infections (GHESKIO), Port-au-Prince, Haiti; University of California, Davis School of Medicine, Davis, California, United States of America; Analysis Group, Boston, Massachusetts, United States of America; Weill Cornell Medical College, New York, New York, United States of America; Brigham and Women’s Hospital, Harvard Medical School, Boston, Massachusetts, United States of America

**Keywords:** HIV, Rapid Antiretroviral Therapy Initiation, Outcome Predictors, Haiti

## Abstract

**Introduction:** Few studies have evaluated baseline predictors of clinical outcomes among people with HIV starting antiretroviral therapy (ART) in the modern era of rapid ART initiation.

**Methods:** We conducted a secondary analysis of a randomized controlled trial of two rapid treatment initiation strategies for people with treatment-naïve HIV and tuberculosis symptoms at an urban clinic in Haiti. We used logistic regression models to assess associations between baseline characteristics and (1) retention in care at 48 weeks, (2) HIV viral load suppression at 48 weeks (among participants who underwent viral load testing), and (3) all-cause mortality.

**Results:** 500 participants were enrolled in the study 11/2017-1/2020. Eighty-eight (18%) participants were diagnosed with tuberculosis, and ART was started in 494 (99%). After adjustment, less than secondary education (adjusted odds ratio [AOR] 0.21, 95% CI 0.10-0.46), dolutegravir initiation (AOR 2.57, 95% CI 1.22-5.43), age (AOR 1.42 per 10-year increase, 95% CI 1.01-1.99), and tuberculosis diagnosis (AOR 3.92, 95% CI 1.36-11.28) were significantly associated with retention. Age (AOR 1.36, 95% CI 1.05-1.75), dolutegravir initiation (AOR 1.75, 95% CI 1.07-2.85), and tuberculosis diagnosis (AOR 0.50, 95% CI 0.28-0.89) were associated with viral suppression. Higher CD4 cell count at enrollment (unadjusted odds ratio [OR] 0.69, 95% CI 0.55-0.87) and anemia (OR 4.86, 95% CI 1.71-13.81) were associated with mortality.

**Conclusions:** We identified sociodemographic, treatment-related, clinical, and laboratory-based predictors of clinical outcomes. These characteristics may serve as markers of sub- populations that could benefit from additional interventions to support treatment success after rapid treatment initiation.

## INTRODUCTION

Rapid initiation of antiretroviral therapy (ART) after HIV diagnosis (i.e., within 7 days of diagnosis, including same-day start) has been recommended by the World Health Organization (WHO) since 2017.^1^ The 2021 WHO guidelines recommend rapid initiation for all people with HIV except for those with concern for meningitis (i.e., secondary to cryptococcus or tuberculosis).^2^ This recommendation was based on multiple randomized controlled trials conducted in diverse settings and published over the last decade.^3–9^ These trials demonstrated that rapid ART initiation results in improvements in retention in care, viral load suppression, and mortality. There have been two hypothesized mechanisms behind this benefit — first, that logistical burdens are minimized; and second, that the more immediate provision of medicine facilitates a sense of hope, optimism, and connection with the health system. While several observational studies using real-world data have suggested that rapid ART initiation may be associated with increased risk of loss to follow-up, these findings can largely be attributed to biases inherent to the study design.^10^

People with newly diagnosed HIV commonly present with signs or symptoms suggesting tuberculosis.^11^ While the 2021 WHO guidelines acknowledged that there is little information available about the risks or benefits of rapid ART initiation among people with HIV and tuberculosis symptoms, they recommended rapid ART initiation while investigating tuberculosis, with close follow-up within seven days to initiate tuberculosis treatment if necessary. Since the publication of these guidelines, our group has demonstrated high rates of ART initiation and retention in care among people with HIV and tuberculosis symptoms in Haiti who were randomized to either of two rapid treatment initiation strategies.^12^

While the recently shifting approach in ART initiation timing reflects the clearly apparent benefits associated with a rapid start, this strategy necessitates a shorter window of time to allow for the assessment of clinical or social risk factors for poor outcomes after initiating ART. As a result, a key research priority in the rapid ART initiation era is to identify readily available baseline characteristics associated with subsequent treatment outcomes such as retention in care, viral suppression, and mortality. A better understanding of these predictors may allow for the development of targeted interventions for sub-populations of people starting ART who are vulnerable to poor outcomes after rapid treatment initiation. To address this gap in the literature, we evaluated baseline predictors of clinical outcomes in a randomized trial of two rapid treatment initiation strategies for people with HIV and tuberculosis symptoms receiving care in urban Haiti.^12^

## METHODS

### Study Design and Oversight

We conducted a secondary analysis of a previously reported open-label randomized controlled trial of treatment initiation strategies for people with treatment-naïve HIV and tuberculosis symptoms in Haiti (NCT03154320).^12^ Our objective in this secondary analysis was to evaluate baseline predictors of (1) retention in care, (2) HIV viral load suppression at 48 weeks, and (3) all-cause mortality. Participants in the trial were randomized 1:1 to either same-day treatment (same-day tuberculosis testing with same-day tuberculosis treatment if diagnosed with tuberculosis; same-day ART if tuberculosis not diagnosed) versus standard care (initiating tuberculosis treatment within seven days and delaying ART to day seven if tuberculosis not diagnosed). For all participants diagnosed with tuberculosis, ART was initiated two weeks after tuberculosis treatment. The trial found no differences by study arm in the primary outcome of retention in care with 48-week HIV-1 RNA <200 copies/mL. Additional details about the treatment strategies and outcomes have previously been reported.^12^ The trial was approved by the institutional review boards at the Haitian Group for the Study of Kaposi’s Sarcoma and Opportunistic Infections (GHESKIO), Mass General Brigham, Florida International University, and Weill Cornell Medical College.

### Setting

The study was conducted at GHESKIO in Port-au-Prince, the urban, densely populated capital of Haiti. During the study there was pervasive and ongoing geopolitical instability, gang violence, and civil unrest in Haiti, with consequent economic instability and hardship. GHESKIO is a Haitian nongovernmental organization and the largest provider of HIV and tuberculosis care in the Caribbean. GHESKIO provides care for approximately 15,000 people with HIV and >2,000 people with tuberculosis annually. The adult HIV prevalence in Haiti is 1.7%, and the annual tuberculosis incidence is 154 per 100,000 persons.^13,14^ First-line treatment for HIV included co-formulated efavirenz, tenofovir disoproxil fumarate, and lamivudine until December 2018, after which dolutegravir became available and was preferred over efavirenz. During 2019, all people with HIV receiving efavirenz-based regimens at GHESKIO were switched to a dolutegravir-based regimen, regardless of viral load. Tuberculosis was treated according to standard guidelines, and all participants who were not diagnosed with tuberculosis disease were treated with prophylactic isoniazid.^15^ Based on recent clinical trial results, participants with CD4 count <100 cells/mm^3^ received azithromycin for five days, and those with CD4 count <100 cells/mm^3^ *and* tuberculosis received prednisone prophylaxis.^16,17^ Retention activities at GHESKIO included: reminder phone calls (or home visits for those without phones) prior to visits, and after missed visits; a transportation subsidy of ∼$1 USD at each visit; and a ∼$1 USD phone card at each visit.

### Study Population

We included all participants enrolled in the original trial in this secondary analysis.^12^ Patients were eligible for inclusion in the trial if they had documented HIV-1 infection, were ≥ 18 years of age, ART-naïve, and reported cough, fever, and/or night sweats of any duration, and/or weight loss. Exclusion criteria included tuberculosis treatment in the past year, lack of preparedness on an ART readiness questionnaire, pregnancy or breastfeeding, active drug or alcohol use, a mental condition that would interfere with completing study requirements, plans to transfer during the study period, symptoms consistent with WHO stage 4 neurologic disease, or WHO “danger signs” of temperature >39 degrees Celsius, pulse >120 beats/minute, respiratory rate >30 breaths/minute, or inability to walk unaided. All participants provided written informed consent.

### Data

Study procedures have been reported in detail elsewhere.^12^ For the purposes of this analysis, we included baseline demographic, clinical and laboratory data, as well as outcome data 48 weeks after enrollment, all of which were extracted from the GHESKIO electronic health record. Baselines variables included in this analysis were: age, sex, income less than $1 USD per day (self-report), educational attainment, marital status, body mass index (BMI), CD4 cell count, tuberculosis diagnosis (microbiologically or clinically diagnosed), and study arm (same-day or standard). We also included information about the ART regimen that was initiated. Outcome variables included clinic attendance at 48 weeks, HIV-1 RNA at 48 weeks, and mortality during 48 weeks of follow-up.

### Outcomes

We considered three outcomes in this analysis. Retention in care was defined as attending a clinical visit 48 weeks after enrollment (with a prespecified window of +/- twelve weeks). Viral load suppression was defined as having HIV-1 RNA<200 copies/mL 48 weeks after enrollment (with a prespecified window of +/- twelve weeks), and was evaluated among all participants who had undergone viral load testing within that window. All-cause mortality over 48 weeks of follow-up after enrollment was determined using family report (death certificates are not available in Haiti).

### Statistical Analysis

Baseline characteristics were summarized using medians and interquartile ranges (IQR) for continuous variables and frequencies and percentages for categorical/binary variables.

We assessed predictors of retention in care, viral suppression, and mortality using logistic regression models. We evaluated associations between our outcome variables and the following baseline predictors: age (continuous variable, per 10 year increase), female sex, income less than $1 USD per day, less than secondary education, married, undernutrition (BMI<18.5 kg/m^2^), CD4 cell count (continuous variable, per 50 cell/mm^3^ increase), anemia (NIH Division of AIDS Grade≥3),^18^ tuberculosis diagnosis, and randomization to the standard strategy study arm. Because first-line ART changed from an efavirenz-based regimen to a dolutegravir-based regimen partway through the study, we also included a binary variable indicating whether a participant had been initiated on dolutegravir. We prespecified this set of 11 predictors and cutoffs for binary predictors from clinical perspectives; we did not conduct model-based variable selection. We did not adjust time-varying covariates that may be less relevant for prediction, or variables that can be affected by study treatment/exposure (e.g., symptoms during follow-up).

We first evaluated univariable associations between predictors and the three outcomes. We then generated multivariable models for the retention in care and viral load suppression outcomes that included all predictors. We did not build a multivariable model for the mortality outcome because there were few deaths (n=15).

Next, for the viral load suppression outcome, we additionally fitted an inverse probability of (censoring) weighting regression model to account for potential selection bias resulting from exclusion of 66 participants who did not undergo viral load testing at week 48.^19,20^ To do this, we weighted observations by the inverse probability that a given participant had a viral load test performed. These probabilities of having non-missing (or complete) data were fitted via multivariable logistic regression that included the 11 baseline covariates in this analysis.^20^

We performed statistical analyses using SAS version 9.4 (SAS Institute, Cary, NC, USA).

## RESULTS

Out of 576 people screened for trial participation between November 2017 and January 2020, 500 were enrolled and are included in this analysis (Table 1).^12^ Median age was 37 years (IQR 30 to 45), and 234 (47%) were female. Cough, fever, night sweats, and weight loss were reported by 200 (47%), 194 (39%), 72 (14%), and 491 (98%) participants, respectively. Median BMI was 20.6 (IQR: 18.7 to 22.9), and 116 (23%) met criteria for undernutrition. Median hemoglobin was 11.8 g/dL (IQR: 10.1, 11.8), and 81 (16%) participants with available hemoglobin results (N=480) had Grade ≥ 3 anemia.^18^ Median CD4 count was 274 cells/mm^3^ (IQR 128 to 426), with 101 (20%) having CD4 count <100 cells/mm^3^. Eighty-eight (18%) participants were diagnosed with tuberculosis at baseline; of these, 68 (77%) were diagnosed microbiologically. All participants with tuberculosis were started on first-line treatment for drug-susceptible tuberculosis.

**Table 1.** Baseline Characteristics of Study Participants. Data are presented as N(%) unless otherwise specified; N=500 unless otherwise specified.

| Characteristics | N (%), unless otherwise specified |
| --- | --- |
| Age at enrollment (years), median (IQR) | 37 (30-45) |
| Female | 234 (47) |
| Income <\$1 USD per day | 372 (74) |
| Less than secondary education | 192 (38) |
| Married | 261 (52) |
| Body mass index (kg/m<sup>2</sup>), median (IQR) | 20.6 (18.7-22.9) |
| Undernutrition (Body mass index <18.5) | 116 (23) |
| CD4 count (cells/mm<sup>3</sup>), median (IQR) (N=495) | 274 (128-426) |
| Hemoglobin (g/dl), median (IQR) (N=480) | 11.8 (10.1-13.4) |
| Anemia (NIH Division of AIDS classification Grade ≥3)* (N=480) | 81 (16) |
| Diagnosed with TB at enrollment | 88 (18) |
| Initiated on dolutegravir-based ART (vs. efavirenz), among ART initiators (N=494) | 184 (37%) |
| Randomized to standard group | 250 (50%) |

\*NIH DAIDS threshold for anemia of Grade ≥3 severity is hemoglobin value ≥9g/dL for males and ≥8.5 g/dL for females.

Antiretroviral therapy was started in 494 (99%) participants — 407 (83%) who were not diagnosed with tuberculosis at baseline, 69 (14%) who initiated ART after initiating tuberculosis treatment, and 18 (4%) who initiated ART before initiating tuberculosis treatment (e.g., in the context of positive culture after initial negative nucleic acid amplification testing). Among participants who started ART, 310 (62%) initiated an efavirenz-based regimen, and 184 (37%) initiated a dolutegravir-based regimen. Among the 412 participants not diagnosed with tuberculosis at baseline, antiretroviral therapy was initiated in 407 (98.8%) at a median time of 6 (IQR 0 to 7) days — 202 (49%) on the day of HIV diagnosis, 110 (27%) within 7 days, 83 (20%) from 8 to 14 days, and 12 (3%) >14 days after HIV diagnosis. Among participants started on efavirenz, 158 (51%) were transitioned to dolutegravir during the study period, a median of 245 days (IQR 201 to 289) after starting ART. Nine participants (2%) who were started on an efavirenz-based regimen were switched to a second-line protease inhibitor-based regimen during the study period.

There were 447 (89%) participants retained in care at week 48, among whom 431 (96%) underwent viral load testing. Among participants who underwent viral load testing, 320 (72%) had HIV-1 RNA <200 copies/mL. Fifteen (3%) participants died during the study period, with causes of death previously reported.^12^

In univariate analyses, less than secondary education (odds ratio [OR] 0.34, 95% CI 0.17 to 0.66) and initiating dolutegravir-based ART (OR 2.74, 95% CI 1.34 to 5.60) were significantly associated with retention in care at 48 weeks (Table 2). After multivariable adjustment, less than secondary education (adjusted odds ratio [AOR] 0.21, 95% CI 0.10 to 0.46) remained significantly associated with a reduced odds of retention in care. Dolutegravir initiation (AOR 2.57, 95% CI 1.22 to 5.43), age (AOR 1.42 per 10 year increase, 95% CI 1.01 to 1.99), and tuberculosis diagnosis at enrollment (AOR 3.92, 95% CI 1.36 to 11.28) were all significantly associated with increased odds of retention.

**Table 2.** Baseline Predictors of Retention in Care at 48 Weeks (N=500)

| Predictors | Univariable Regression |  | Multivariable Regression** |  |
| --- | --- | --- | --- | --- |
|  | OR (95% CI) | p-value | OR (95% CI) | p-value |
| Age (per 10 year increase) | 1.19 (0.88, 1.61) | 0.25 | 1.42 (1.01, 1.99) | 0.04 |
| Female sex | 0.98 (0.56, 1.74) | 0.95 | 1.28 (0.67, 2.43) | 0.46 |
| Income <\$1 USD per day | 0.84 (0.42, 1.64) | 0.60 | 0.80 (0.37, 1.70) | 0.55 |
| Less than secondary education | 0.34 (0.17, 0.66) | 0.001 | 0.21 (0.10, 0.46) | <.0001 |
| Married | 0.82 (0.46, 1.46) | 0.50 | 0.72 (0.38, 1.35) | 0.30 |
| Undernutrition | 1.04 (0.53, 2.04) | 0.92 | 0.97 (0.46, 2.05) | 0.93 |
| CD4 count at enrollment (per 50 cells/mm<sup>3</sup> increase) | 1.01 (0.95, 1.08) | 0.67 | 1.04 (0.97, 1.10) | 0.30 |
| Anemia (Grade ≥3)* | 0.81 (0.39, 1.69) | 0.58 | 0.51 (0.23, 1.16) | 0.11 |
| Diagnosed with tuberculosis at enrollment | 2.19 (0.85, 5.67) | 0.11 | 3.92 (1.36, 11.28) | 0.01 |
| Randomized to standard group (vs. same-day group) | 1.60 (0.90, 2.87) | 0.11 | 1.73 (0.93, 3.25) | 0.09 |
| Dolutegravir-based ART | 2.74 (1.34, 5.60) | 0.006 | 2.57 (1.22, 5.43) | 0.01 |

\*NIH DAIDS threshold for anemia of Grade ≥3 severity is hemoglobin value ≥9g/dL for males and ≥8.5 g/dL for females.
\*\* Missing data were present in 5 persons. AUC=0.75.

Among participants who underwent viral load testing at 48 weeks, age (OR 1.35 per 10 year increase, 95% CI 1.08 to 1.71), tuberculosis diagnosis (OR 0.42, 95% CI 0.25 to 0.69), and dolutegravir initiation (OR 1.81, 95% CI 1.13 to 2.90) were significantly associated with viral suppression (Table 3). After multivariable adjustment, age (AOR 1.36, 95% CI 1.05 to 1.75) and dolutegravir initiation (AOR 1.75, 95% CI 1.07 to 2.85) remained positively associated with viral suppression, and tuberculosis diagnosis (AOR 0.50, 95% CI 0.28 to 0.89) remained negatively associated with viral suppression. Findings were similar after incorporating inverse probability weights to account for participants who did not undergo viral load testing (Table 3). Weights ranged from 1.02 to 1.48, which means 1 complete case/person represented approximately 1 to 1.5 persons/observations from the overall population.

**Table 3.** Baseline Predictors of Viral Suppression among Patients who Completed 48-week HIV-1 RNA Testing.

| Predictors | Univariable Analysis<br>(N=431) |  | Multivariable Analysis<br>(N=429)** |  | Multivariable Analysis with<br>Inverse Probability Weights<br>(N=429)*** |  |
| --- | --- | --- | --- | --- | --- | --- |
|  | OR (95% CI) | p-value | OR (95% CI) | p-value | OR (95% CI) | p-value |
| Age (per 10 year increase) | 1.35 (1.08, 1.71) | 0.01 | 1.36 (1.05, 1.75) | 0.02 | 1.35 (1.07, 1.70) | 0.01 |
| Female sex | 1.05 (0.68, 1.62) | 0.82 | 1.05 (0.66, 1.67) | 0.84 | 1.05 (0.68, 1.62) | 0.83 |
| Income <\$1 USD per day | 0.90 (0.55, 1.49) | 0.69 | 1.11 (0.64, 1.90) | 0.72 | 1.10 (0.66, 1.82) | 0.72 |
| Less than secondary education | 0.89 (0.57, 1.37) | 0.58 | 0.78 (0.48, 1.27) | 0.32 | 0.79 (0.50, 1.23) | 0.29 |
| Married | 1.44 (0.93, 2.22) | 0.10 | 1.42 (0.90, 2.26) | 0.14 | 1.49 (0.97, 2.29) | 0.07 |
| Undernutrition | 0.70 (0.43, 1.14) | 0.15 | 0.89 (0.25, 1.54) | 0.68 | 0.87 (0.52, 1.44) | 0.58 |
| CD4 count at enrollment (per 50 cells/mm<sup>3</sup> increase) | 1.04 (0.99, 1.09) | 0.16 | 1.03 (0.98, 1.08) | 0.31 | 1.03 (0.98, 1.08) | 0.28 |
| Anemia (Grade ≥3)* | 0.82 (0.46, 1.46) | 0.50 | 1.07 (0.57, 2.01) | 0.83 | 1.05 (0.58, 1.90) | 0.87 |
| Diagnosed with tuberculosis at enrollment | 0.42 (0.25, 0.69) | 0.0008 | 0.50 (0.28, 0.89) | 0.02 | 0.50 (0.29, 0.86) | 0.01 |
| Randomized to standard group (vs. same-day group) | 1.25 (0.81, 1.93) | 0.31 | 1.15 (0.73, 1.81) | 0.54 | 1.11 (0.73, 1.69) | 0.63 |
| Dolutegravir-based ART | 1.81 (1.13, 2.90) | 0.01 | 1.75 (1.07, 2.85) | 0.03 | 1.72 (1.09, 2.72) | 0.02 |

\*NIH DAIDS threshold for anemia of Grade ≥3 severity is hemoglobin value ≥9g/dL for males and ≥8.5 g/dL for females
\*\* Missing data were present in 2 persons. AUC=0.66.
\*\*\* Missing data were present in 2 persons. AUC=0.66. CIs were estimated assuming weight is known since very similar
CIs were obtained when weight is known or estimated.

Higher CD4 cell count at enrollment was significantly associated with a lower odds of mortality in unadjusted models (OR 0.69, 95% CI 0.55 to 0.87) (Table 4). Grade 3 ≥ anemia was associated with a significantly greater odds of mortality (OR 4.86, 95% CI 1.71 to 13.81).

**Table 4.** Baseline Predictors of Mortality.

| Predictors | Univariable Analysis* |  |
| --- | --- | --- |
|  | OR (95% CI) | p-value |
| Age (per 10 year increase) | 1.25 (0.76, 2.06) | 0.38 |
| Female sex | 0.99 (0.36, 2.79) | 0.99 |
| Income <\$1 USD per day | 0.50 (0.18, 1.45) | 0.20 |
| Less than secondary education | 1.59 (0.54, 4.73) | 0.40 |
| Married | 0.60 (0.21, 1.72) | 0.34 |
| Undernutrition | 1.69 (0.56, 5.03) | 0.35 |
| CD4 count at enrollment (per 50 cells/mm<sup>3</sup> increase) | 0.69 (0.55, 0.87) | 0.002 |
| Grade ≥3 anemia* | 4.86 (1.71, 13.81) | 0.003 |
| Diagnosed with tuberculosis at enrollment | 0.33 (0.04, 2.52) | 0.28 |
| Randomized to standard group (vs. same-day group) | 0.66 (0.23, 1.88) | 0.43 |
| Initiated on dolutegravir-based ART | 0.62 (0.19, 1.96) | 0.41 |

\*We did not conduct Multivariable regression due to only 15 deaths.

## DISCUSSION

In this study of 500 people with newly diagnosed HIV and tuberculosis symptoms (about one in five of whom were diagnosed with tuberculosis) who participated in a randomized trial of two rapid treatment initiation protocols, we identified sociodemographic (age, educational attainment), treatment-related (dolutegravir-based regimens), clinical (tuberculosis diagnosis), and laboratory-based (CD4 count and anemia) predictors of clinical outcomes over a follow-up period of 48 weeks. Despite substantial geopolitical instability in Haiti during the study period, nearly all participants initiated ART (over three quarters of participants without tuberculosis started within 7 days) and retention in care was high. This is one of the first studies to report baseline predictors of clinical outcomes in the context of rapid ART initiation, especially among people experiencing tuberculosis symptoms. The predictors we identified can be easily and quickly measured during the condensed evaluation prior to rapid ART initiation, and may serve as markers of sub-populations that could benefit from additional interventions to support treatment success.

The two sociodemographic factors associated with poorer outcomes were younger age (associated with lower retention in care and lower viral suppression rates) and less than a secondary education (associated with lower retention in care). Both of these factors have previously been identified as predictors of clinical outcomes in pre-rapid ART initiation settings,^21^ and our findings demonstrate that they continue to be important markers in the current era.

Education was the baseline characteristic most strongly associated with retention in care. Education was also identified as a key driver of persistent disparities in life expectancy among patients initiating ART at GHESKIO during earlier time periods.^22^ Our results may reflect several potential mechanisms. First, lower education may have been associated with lower health literacy, with greater consequent difficulty communicating the benefits of ART and clinical follow-up. If this is the case, patients with lower education levels may benefit from tools designed to facilitate effective and rapid communication of key HIV-related concepts.^23,24^ While this may be exacerbated by the shorter duration of pre-treatment counseling inherent to rapid ART initiation, as we noted above this association was also described prior to the adoption of rapid ART initiation.^21^ Second, this association may result from an unmeasured social or economic factor that is correlated with education. For example, food insecurity tends to be strongly associated with educational attainment, is highly prevalent in Haiti, and has been associated with a wide variety of adverse HIV outcomes.^25–28^ More generally, socioeconomic factors like poverty are associated with education and can strongly influence HIV outcomes.^29^ While future research should aim to disentangle these potential mechanisms, educational level can be used in HIV programs as a prognostic marker for clinical outcomes after treatment initiation.

Initiation of a dolutegravir-based regimen (versus an efavirenz-based regimen) was associated with an increased odds of both retention in care and viral suppression. This finding further supports the preference for dolutegravir as first-line therapy,^2^ is consistent with dolutegravir’s greater tolerability and effectiveness relative to efavirenz,^30^ and illustrates the instrumental role that dolutegravir can play in improving HIV-related population health. This is especially the case in settings with high rates of efavirenz resistance among ART-naïve people with HIV, like Haiti.^31^ Of note, about half the participants started on a efavirenz-based ART were transitioned to dolutegravir at some point during the study period. This likely biased the association between dolutegravir initiation and clinical outcomes towards the null, meaning that the benefits of dolutegravir are likely to be even greater than those estimated in this study.

While all participants in this study were experiencing tuberculosis symptoms at enrollment, only 18% were ultimately diagnosed with tuberculosis, and being diagnosed with tuberculosis was significantly associated with greater odds of retention in care and lower odds of viral suppression. Greater retention likely results from closer clinical contact during tuberculosis treatment (e.g., directly observed therapy) and the fact that people with tuberculosis commonly experience rapid improvement in symptoms on therapy. Other studies have also reported an association between virologic failure and tuberculosis co-infection.^32–35^ Lower odds of viral suppression after tuberculosis diagnosis may relate to delays in ART initiation (although these tended to be short), difficulty with drug tolerability given additional medications being administered. Tuberculosis may also increase HIV viral load and the size of the HIV reservoir.

There were few deaths among participants during follow-up, and two baseline laboratory characteristics were significantly associated with death — CD4 cell count and anemia. Immunologic dysfunction from progressive HIV has long been recognized as a key risk factor for death among people with HIV. Anemia, which may result from both nutritional and non-nutritional (e.g., disseminated infection) mechanisms, has also been identified as an important risk factor for mortality.^36,37^

This study had several limitations. It was conducted among participants reporting tuberculosis symptoms at a single large urban clinic with a high quality of care, which may limit generalizability to other settings and contexts. Our viral suppression outcome was assessed among participants who completed viral load testing at week 48, which raises the possibility of selection bias. However, over 85% of participants underwent viral load testing, and estimates from our missing data-adjusted analyses were nearly identical to those from an unweighted multivariable model. Lastly, the odds ratios reported here are associational, not casual, with non-randomized exposures, which is typical in studies of identification of predictors or risk or protective factors.

## CONCLUSIONS

In this study of 500 people with newly diagnosed HIV and tuberculosis symptoms (18% of whom were diagnosed with tuberculosis) who participated in a trial of two rapid treatment initiation protocols, we identified sociodemographic (age, educational attainment), treatment-related (dolutegravir-based regimens), clinical (tuberculosis diagnosis), and laboratory-based (CD4 count and anemia) predictors of clinical outcomes over a follow-up period of 48 weeks. This is one of the first studies to report baseline predictors of clinical outcomes in the context of rapid ART initiation, especially among people reporting tuberculosis symptoms. The predictors we identified can be easily and quickly assessed during the condensed evaluation prior to rapid ART initiation, and may serve as markers of sub-populations that could benefit from additional interventions to support treatment success.

## DECLARATIONS

All authors declare no conflicts of interest.

## Data Availability

A deidentified dataset is available online at https://journals.plos.org/plosmedicine/article?id=10.1371/journal.pmed.1004246

